# Combining Transfer Learning with Retinal Lesions Features for Accurate Detection of Diabetic Retinopathy

**DOI:** 10.1101/2022.09.23.22280273

**Authors:** Doaa Hassan, Hunter Mathias Gill, Michael Happe, Ashay D. Bhatwadekar, Amir R. Hajrasouliha, Sarath Chandra Janga

**Affiliations:** Department of BioHealth Informatics, School of Informatics and Computing, Indiana University Purdue University, 535 West Michigan Street, Indianapolis, Indiana 46202; Department of Ophthalmology, Glick Eye Institute, Indiana University School of Medicine, e, 1160 W. Michigan Street Indianapolis, IN 46202; Department of Medical and Molecular Genetics, Indiana University School of Medicine, Medical Research and Library Building, 975 West Walnut Street, Indianapolis, Indiana, 46202; Centre for Computational Biology and Bioinformatics, Indiana University School of Medicine, 5021 Health Information and Translational Sciences (HITS), 410 West 10th Street, Indianapolis, Indiana, 46202; Computers and Systems Department, National Telecommunication Institute, Cairo, Egypt

**Keywords:** retinal image, diabetic retinopathy, deep learning, transfer learning, lesion features

## Abstract

Diabetic retinopathy (DR) is a late microvascular complication of Diabetes Mellitus (DM) that could lead to permanent blindness in patients, without early detection. Although adequate management of DM via regular eye examination can preserve vision in in 98% of the DR cases, DR screening and diagnoses based on clinical lesion features devised by expert clinicians; are costly, time-consuming and not sufficiently accurate. This raises the requirements for Artificial Intelligent (AI) systems which can accurately detect DR automatically and thus preventing DR before affecting vision. Hence, such systems can help clinician experts in certain cases and aid ophthalmologists in rapid diagnoses. To address such requirements, several approaches have been proposed in the literature that use Machine Learning (ML) and Deep Learning (DL) techniques to develop such systems. However, these approaches ignore the highly valuable clinical lesion features that could contribute significantly to the accurate detection of DR. Therefore, in this study we introduce a framework called DR-detector that employs the Extreme Gradient Boosting (XGBoost) ML model trained via the combination of the features extracted by the pretrained convolutional neural networks commonly known as transfer learning (TL) models and the clinical retinal lesion features for accurate detection of DR. The retinal lesion features are extracted via image segmentation technique using the UNET DL model and captures exudates (EXs), microaneurysms (MAs), and hemorrhages (HEMs) that are relevant lesions for DR detection. The feature combination approach implemented in DR-detector has been applied to two common TL models in the literature namely VGG-16 and ResNet-50. We trained the DR-detector model using a training dataset comprising of 1840 color fundus images collected from e-ophtha, retinal lesions and APTOS 2019 Kaggle datasets of which 920 images are healthy. To validate the DR-detector model, we test the model on external dataset that consists of 81 healthy images collected from High-Resolution Fundus (HRF) dataset and MESSIDOR-2 datasets and 81 images with DR signs collected from Indian Diabetic Retinopathy Image Dataset (IDRID) dataset annotated for DR by expert. The experimental results show that the DR-detector model achieves a testing accuracy of 100% in detecting DR after training it with the combination of ResNet-50 and lesion features and 99.38% accuracy after training it with the combination of VGG-16 and lesion features. More importantly, the results also show a higher contribution of specific lesion features toward the performance of the DR-detector model. For instance, using only the hemorrhages feature to train the model, our model achieves an accuracy of 99.38 in detecting DR, which is higher than the accuracy when training the model with the combination of all lesion features (89%) and equal to the accuracy when training the model with the combination of all lesions and VGG-16 features together. This highlights the possibility of using only the clinical features, such as lesions that are clinically interpretable, to build the next generation of robust artificial intelligence (AI) systems with great clinical interpretability for DR detection. The code of the DR-detector framework is available on GitHub at https://github.com/Janga-Lab/DR-detector and can be readily employed for detecting DR from retinal image datasets.

## 1. Introduction

Diabetic Retinopathy (DR) is a microvascular disorder associated with long-term diabetes mellitus and is one of the leading causes of preventable vision loss across the worldwide [1]. DR manifests in individuals diagnosed with Type 1 Diabetes (T1D) or Type 2 Diabetes (T2D). Roughly one-third of diabetic patients are affected by DR [2,3], and the likelihood of developing DR scales with the length of diabetes duration [4].

The progression of DR in T1D and T2D is characterized by damage to the retina. The retina is a multilayered network of rod and cone photoreceptor cells integrated with bipolar and ganglion cells that enable vision by encoding information gained from light as nerve impulses [5]. The retina is supplied with oxygen and nutrients by an extensive vascular system. In T1D and T2D, high blood glucose levels contribute to pro-inflammatory changes that increase the permeability of the blood-retina barrier, leading to leakage of fluids and blood into the retina [6]. High blood glucose can also block small retinal capillaries, impeding the delivery of nutrients and contributing to further damage [7].

Although adequate management of DM via regular eye examination can preserve vision in DR in many cases, DR screening and diagnoses currently involve highly trained and qualified medical professionals at a high cost. Thus, there is a continuous need for the development of automatic approaches for DR detection as a cheaper alternative to the time-consuming manual DR diagnosis by trained clinicians. A promising application of these approaches is Computer Assisted Diagnosis (CAD) support for detection of DR. An advantage of such CAD applications is that they offset the burden on medical professionals like expert ophthalmologists and fill their absence in addition to preventing DR before affecting vision. This consideration is critical, considering that the global burden of DR is expected to expand to 700 million cases by the 2040s [8]. Many DR detection techniques suitable for CAD utilize Machine Learning (ML), Deep Learning (DL) algorithms and various previously pretrained DL models commonly known as Transfer Learning (TL) models.

The TL models have been successfully used for automated binary and multi-class classification of color fundus retinal images for DR detection [9–12]. These algorithms have shown a great performance in the automatic detection of DR in non-clinical setups when the dataset is very small and might cause chances for underfitting or high generalization error. In such cases, TL is preferred over standard DL techniques. Recently, the focus has been shifted to TL feature-based models, where common TL algorithms are used for extracting many important local (textural) features from retinal images for detection of DR and predicting its severity level through convolving with a sliding window and forming a filter. For example, features extracted from AlexNet TL model [13] were passed to the Support Vector Machine (SVM) ML model to enhance the efficiency of the DR classification system, where SVM model achieved accuracies of 97.93% and 95.26% in five-class DR classification with linear discriminant analysis (LDA) feature selection and principle component analysis (PCA) dimensional reduction respectively [14], when training the model and testing on Kaggle dataset^1^. As an extension to the same direction, features from the final layers of VGG-19 TL model [15] were collected and aggregated to get a deeper representation of retinal images, and these dense features were reduced by PCA and singular value decomposition (SVD) [16], where it was fed to a deep Neural network (DNN) model that achieved accuracies of 97.96%, and 98. 34% in DR severity classification with PCA and SVD respectively when training the model and testing on Kaggle dataset. M.K.Yaqoob et al.[17] introduced a feature representation extracted by ResNet-50 TL model [18] that was fed to Random Forest (RF) classifier for binary and multiclassification of DR. This approach aheived an accuracy of 96% when it was applied on a dataset comprised of two DR categories for detecting DR and accuracy of 75.09% when it was applied as on five DR category dataset for predicting the severity of DR.

Bodapati JD et al. [19] introduced a DR classification model that aggregates features extracted from multiple convolution blocks of TL models to enhance feature representation and hence improve DR detection. The model has compared various methodologies of pooling and feature aggregation and it was concluded that averaging pooling with simple fusion approaches using Deep Neural Networks (DNN) led to an improved performance. The authors of this work claimed that their approach for blending features from the convolution layers of the same TL model is simpler and better than the simple concatenation of features extracted from various TL models at a different scale that was presented early in [20]. The latter approach introduced a multi-modal blended TL feature representation for extracting deep features from penultimate layers of multiple TL models and blending them using different pooling approaches to obtain the final DR image representation.

However, besides the local features presented by various TL models or the fusion of those features, the global image features have been playing an important role in DR detection. Those features are represented by the contour and structural features that describe retinal lesions like exudates (EXs), microaneurysms (MAs), and hemorrhages (HEMs), where the presence of DR disease is characterized by detecting one or more of these lesions. Thus, those global features (lesion features) are considered as good signs of retinal image lesions and hence can be successfully applied to improve the final accuracy of a TL-feature based DR screening system. Therefore, there were some attempts that used image segmentation techniques for extracting/detecting retinal lesion features that can be used for DR detection and staging [21,22]. In such image segmentation-based methods, a label is assigned to every pixel of an image based on pixel characteristics [23]. The labels are encoded in a segmentation mask with equal dimensions to the image. In binary segmentation tasks, each mask pixel represents either the foreground (corresponds to an area-of-interest in the image; value = 1) or the background (corresponds to all non-area-of-interest; value = 0). Thus, binary segmentation tasks are useful for extracting notable areas from biomedical images.

With the introduction of deep learning (DL), especially convolutional neural network (CNN), the DL based methods have resulted in an outstanding performance in medical image segmentation [24]. UNET-architecture CNN models are one of DL models that achieve remarkable performance in medical image segmentation tasks [25]. For the task of retinal image segmentation for DR detection, there were multiple research reports that demonstrate the use of UNET for segmentation of leakage-prone blood vessels [26,27]. UNET models have also been successfully developed for the segmentation of MAs [28–31], EXs [32–35] and HEMs [36].

Although TL and segmentation features provide robust information for DR detection, they were not used together in the literature for a two-class (binary) DR-detection. Only a few existing research attempts utilized the fusion of both types of features for improving the performance of predicting the severity levels of DR [37,38]. For example, B. Harangi et al. proposed a framework that combines AlexNet TL-based features with image-level features that reflect the intensity, shape, and texture of the structures of the image and lesion-specific features associated with MAs and EXs. This combination of features was passed through an additional fully connected layer followed by a softmax function that achieved an accuracy of 90.07% in predicting the class probabilities corresponding to 5 classes for DR that express its severity levels. In [38], the same idea was extended to several commonly used TL models for local image feature extraction other than AlexNet. Next, the results of concatenating the TL features of those models with the hand-crafted features (image level and lesion features) were objectively compared to demonstrate the best concatenation framework that improves the accuracy of predicting DR grades. Next, the best concatenation was passed through an additional fully connected layer then a softmax function to predict the five class probabilities of DR severty. However, both approaches in [37,38] were not tested for binary classification of DR to report the presence of the disease. Also, the performance of the lesion feature extraction was not explicitly investigated as well as the impact of those features on DR detection. Moreover, both approaches combined the image level features with TL and lesion features for training the DR severity levels predictors, increasing the curse of dimensionality.

Therefore, in this study we propose a framework called DR-detector (Figure 1) that employs the XGboost ML model [39] for an accurate detection of DR. The model is trained with a combination of the TL features, and three clinical lesion features that capture EXs, MAs, and HEMs. Such a combination is used to get a better representation of retinal image features that can be used to decide about the presence of DR disease. Thus, we seek the power of TL model to extract features that accurately capture the local textural retinal images while simultaneously taking advantage of the power of lesion features to represent the global features of the retinal images that would result in improving the performance of the DR-detector model and its interpretability for clinical use. We tested the DR-detector model on an external dataset of retinal images for detection of DR. We have applied our proposed framework to two common TL models in the literature namely VGG-16 and ResNet-50 models (Figure 1A). The experimental results show that the DR-detector model achieves an accuracy of 100% in detecting DR when testing it on an external dataset after training it with the combination of Resnt-50 and lesion features and 99.38% accuracy after training it with the combination of VGG-16 and lesion features. The results also show a higher contribution of some lesion features towards the performance of the model over other lesion features. For instance, using the hemorrhages feature to train the model, our model achieves an accuracy of 99.38 in detecting DR which is higher than the accuracy of the model when training it with the combination of all lesion features (89%) and equal to the accuracy when training it with the combination of all lesions and VGG-16 features together. Thus, we arrived at two major conclusions. First, the extracted relevant lesion features can complement the textural features extracted by the TL model to improve the performance of DR-detector model and its interpretability for clinical use in detecting the presence of DR disease. Second, the contribution of lesion features to the performance of DR-detector model varies from one lesion feature to another. This highlights the possibility of using only the lesion features for training the next generation of robust and accurate AI models with clinical interpretability for DR detection.

**Figure 1.**
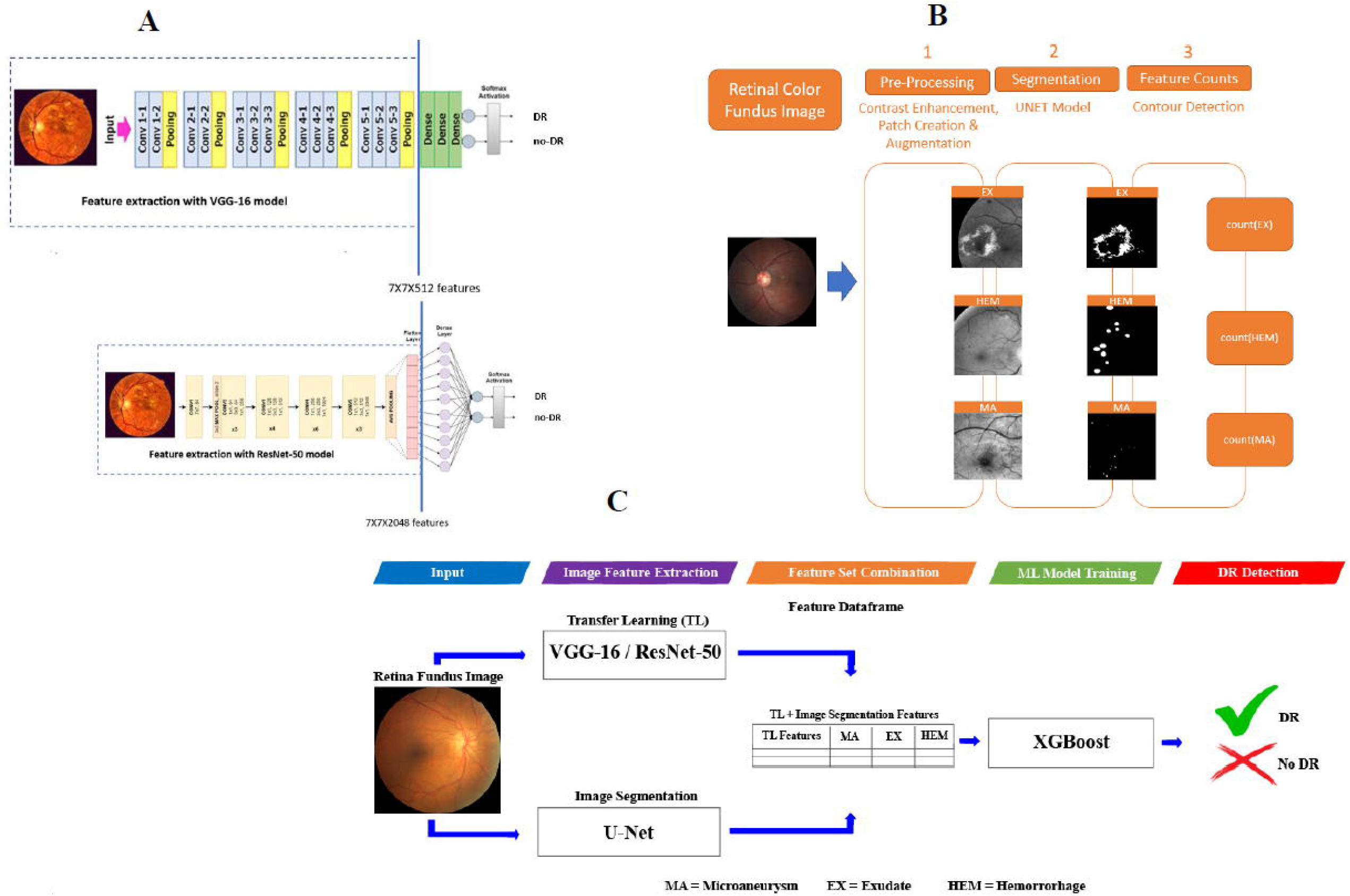
The workflow of DR-detector framework for detection of DR using a combination of TL and lesion features

## 2. Material and Methods

### 2.1 Approach pipeline

The main objective of this work is to develop a robust and efficient framework called DR-detector for automatic detection of DR. Thus, we employed ML model namely the XGBoost in this framework to achieve this objective. This model is trained with a combination of two types of extracted features. The first type is deep convolutional features extracted using a TL model (VGG16-model or Resnet50) pre-trained previously on ImageNet dataset [40] (Figure 1A). Those deep features were known as the most descriptive and discriminate features that ultimately improve the performance of DR recognition [16]. The second type of features are three clinical lesion features that capture the EXs, MAs, and HEMs and are extracted using image segmentation via U-Net DL model (Figure 1B). Those lesion features were found to be the most common pathological signs of DR in the literature [41]. Next the performance of DR-detector model is tested on external dataset of fundus retinal images after training it with the combination of TL and lesion features (Figure 1C). In summary, the proposed pipeline of the DR-detector framework (Figure 1) has five different modules including TL feature extraction (Figure 1A), lesion feature extraction (Figure 1B), and feature combination, model training and model evaluation (Figure 1C).

### 2.2 Datasets

#### 2.2.1 Training dataset

We conduct our experiments on 1840 color fundus images. 920 images of them have DR signs, and the remaining are healthy images. Those images were used for training the DR-detector model. The images with DR signs were collected from two public-available datasets, namely the e-ophtha [42], and retinal lesions [43,44] with binary masks for extracting and quantifying the EXs, MAs and HEMs lesion features. The healthy images were collected from APTOS 2019 Blindness Detection Kaggle competition training dataset [45]. The Binary masks for healthy eye images are generated by creating all-black images with identical dimensions.

The images in the training dataset have various levels of DR on a scale of 0 to 4 (0 - No DR (healthy), 1-Mild, 2-Moderate, 3-Severe, 4-Proliferative DR) to indicate different DR severity levels. However, the data is imbalanced as it consists of 920 healthy images of DR-level 0, 120 images of DR-level 1, 681 images of DR-level 2, 64 images of DR level 3, 55 images of DR level 4. Since we found that there is not enough data from DR classes of DR-levels (1-4) that we can include in the training dataset to balance it, we decided to go with the binary classification of

DR for automatic detection of this disease. Therefore, we adopt the training data set for binary classification problem by merging all images of DR signs of 1-4 into a single positive class of 920 images labeled as DR and the remaining 920 images are labeled as healthy and assigned to the negative class as shown in Table 1.

**Table 1:** Number of Healthy and DR images in the training dataset

| DR severity level | Number of healthy and DR images |
| --- | --- |
| 0 | 920 (Healthy) |
| 1 | 920 (DR) |
| 2 |  |
| 3 |  |
| 4 |  |
| Total | 1840 |

#### 2.2.2 Testing dataset

For testing the DR-detector model, we have used a dataset of 162 color fundus images, where 81 of them are annotated as DR affected, and the remaining are from healthy individuals. Those images were collected from three publicly available datasets, namely High-Resolution Fundus (HRF) dataset [46], Indian Diabetic Retinopathy Image Dataset (IDRID) dataset [47], and MESSIDOR-2 datasets [48]. The IDRID contained 81 color fundus images (4288 × 2848) with binary masks representing DR-affected eyes needed to extract and quantify the EXs, MAs and HEMs lesion features. However, IDRID dataset does not contain any healthy eye images, so the healthy eye images in the testing dataset were randomly selected from HRF and MESSIDOR-2 datasets. Binary masks for healthy eye images are generated by creating all-black images with identical dimensions.

Similar to the training data, the images in the testing dataset have 0-4 levels of DR to indicate different DR severity levels. However, the dataset is imbalanced as it consists of 81 healthy images of DR-level 0 and 81 images of DR affected images with 2 images of DR-level 1, 34 images of DR-level 2, 22 images of DR level 3, and 23 images of DR level 4. Since there is not enough data from DR classes of DR-levels (1-4) that we can include in the testing dataset to balance it, it has been more convenient to use such data for binary classification of DR for detection of the disease. To achieve this, we adopt the testing data set for binary classification problem by merging all images of DR levels of 1-4 into a single positive class of 81 images labeled as DR and the remaining 81 images are labeled as healthy and assigned to the negative class as shown in Table 2.

**Table 2:** Number of Healthy and DR images in the testing dataset

| DR severity level | Number of healthy and DR images |
| --- | --- |
| 0 | 81 (healthy) |
| 1 | 81 (DR) |
| 2 |  |
| 3 |  |
| 4 |  |
| Total | 162 |

### 2.3 Image feature extraction with Transfer Learning

In this approach, local representations of the retinal image’s features are obtained from the TL model (either the VGG16 or the Resent50 pretrained models) by extracting deep features from the final layers of the pre-trained models. When performing feature extraction with TL models, we treat the pre-trained network as an arbitrary feature extractor, allowing the input image to propagate forward, stopping at pre-specified layer, and taking the outputs of that layer as our features.

As for extracting deep features using VGG-16 pretrained model, the original VGG-16 model [15] is adopted first to address the automatic detection of DR (top subfigure of Figure 1A). For this task, the model expects input images of 224*224*3. Thus, images are reshaped to 224*224*3 before feeding them to this model. Next the soft-max layer and fully connected (FC) layers are removed from VGG-16 model (area after the solid vertical blue line in top subfigure of Figure 1A and the model utilizes the VGG-16 network [15] for feature extraction via the final layer prior to the FC layers — that outputs volume of size 7 × 7 × 512 dim (area with dashed border in top subfigure of Figure 1A). This output will serve as VGG-16 extracted features which will be flattened later into a feature vector of 25,088-dim combined with the lesion features, as described later in section 2.5.

As for extracting deep features using the ResNet-50 pretrained model, the original ResNet-50 model [17] is adopted first to address the detection of DR task (bottom subfigure of Figure 1A). For this task, the model expects input images of 224*224*3. Thus, images are reshaped to 224*224*3 before feeding them to this model. Next, the soft-max layer and fully connected (FC) layers are removed from ResNet-50 model (area after the solid vertical blue line in the bottom subfigure of Figure 1A) and the model utilizes the ResNet-50 network [18] for feature extraction via the final layer before the fully connected (FC) layers — that outputs volume of size 7 × 7 × 2048 dim (area with dash border in bottom subfigure of Figure 1A). This output will serve as ResNet-50 extracted features which will be flattened later into a feature vector of 100,352-dim combined with lesion features as described later in section 2.5.

### 2.3 lesion feature extraction with image segmentation

#### 2.4.1 Retinal image lesions associated with DR

Retinal lesions that develop early over the course of DR, including MAs, EXs, and HEMs (Supplementary Figure 1), are clinically important markers used to distinguish between healthy and DR-affected eyes. Below, we elaborate about each of these three lesions:

##### Microaneurysms

MA are the earliest symptoms of DR. These lesions are widened protrusions extending from capillary walls and are associated with abnormal fluid leakage through breakdown of the blood-retina barrier. MA can rupture to create hemorrhages, leading to greater leakage of capillary fluids and damage to surrounding retinal tissues. The number of microaneurysms can be used to gauge the progression of DR [49].

##### Exudates

EXs are lipids and proteins (fibrinogen, albumin) carried by exfiltrating fluids past the blood-retina barrier into the retinal tissue [50]

##### Hemorrhages

HEMs occur when MA burst, and leak blood and serum into the retina. Intraretinal bleeding is a sign of worsening DR. Blood can impair DR patient vision and increased intraretinal pressure can contribute to retinal damage. [51]

#### 2.4.2 Framework for UNET-model-based Lesion Detection & Quantification

We have developed a framework to extract MAs, EXs, and HEMs lesions from retinal fundus images using U-Net semantic segmentation models (Figure 1B) and deployed it in DR-detector framework for extracting lesion features. The steps for the UNET-based retinal lesion detection and quantification workflow are described below:

##### Preprocessing

Binary thresholding is applied to set all pixels corresponding to the image background (the non-eye margins) equal to zero. Multiple studies demonstrate the green channel encodes the greatest contrast between retinal structures [32–34]. Input RGB retinal fundus images are split by channel and the green channel is extracted. Contrast Limited Adaptive Histogram Equalization (CLAHE) (8×8 tile size) [52] is applied to the green channel to correct the contrast over-amplification. A gamma correction is utilized to adjust luminescence of the CLAHE output (γ = 0.8). The contrast enhancement stages are shown in Supplementary Figure 2. After contrast enhancement, images are divided into patches.

**Figure 2.**
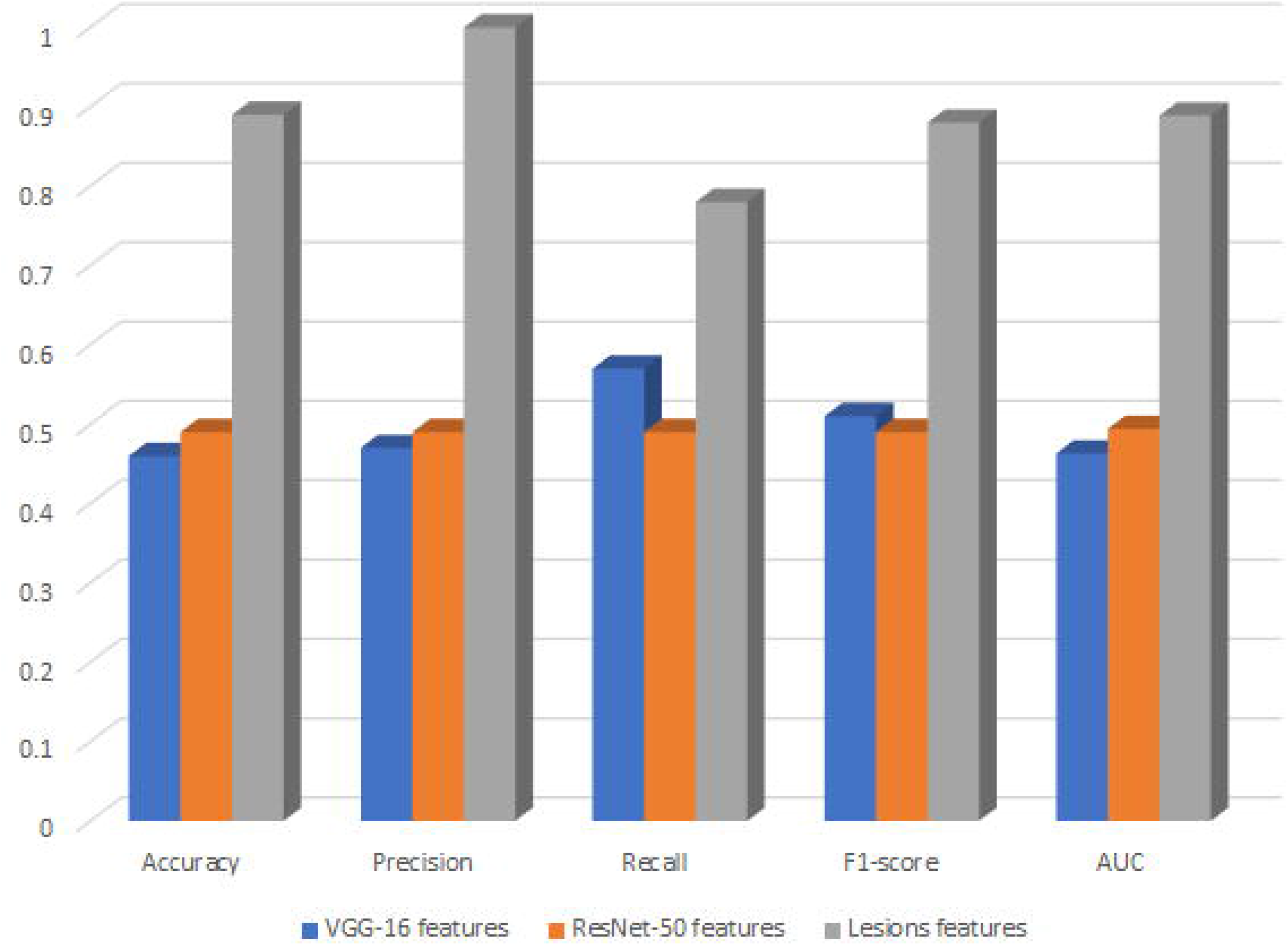
The performance of XGBoost with VGG-16, ResNet-50 and lesions features.

Each preprocessed retinal image and its corresponding ground truth mask is divided into overlapping square (*n* x *n)* patches. *n* is set to 128 pixels (px) for MAs (due to small lesion size) and is set to 256 px for EXs and HEMs. Created patches are randomly selected for augmentation operations.

Augmentation for image and corresponding binary mask patches involves creating new training instances from existing ones by applying a spatial or color operation to represent them in a new orientation or perspective. The random flip (horizontal, vertical) and random rotation (360°) techniques from Keras ImageDataGenerator [53] are used to augment training patches. Any augmentation technique applied to a fundus image patch is likewise applied to its ground truth patch.

##### Segmentation

The input retinal fundus images are preprocessed and divided into augmented patches as described above. K-fold cross-validation (k = 5) is applied to the patches. The batch size for each fold is set to 32 and the number of epochs is set to 3. Epoch training and validation steps are set as the number of training or validation patches per fold divided by batch size, respectively.

Patch probability maps output by the UNET DL model are merged to construct a probability map with equal dimensions to the input image. A threshold of 0.5 is applied to convert the reconstructed probability map into a binary image mask.

##### Feature Counts

Canny edge detection [54] is applied to find the edges around mask foreground regions (white). Contour detection [55] is used to fill Canny edge gaps and fully close the foreground shapes. The number of lesions within the segmentation mask is defined as the number of distinct objects described by contours.

#### 2.4.3 U-Net model implementation

Keras [56], the free python deep learning API with TensorFlow [57] back end was used to construct a base UNET model for binary semantic segmentation. Input patches are supplied to the input layer as tensors with shape (32, *n, n*, 1). The UNET contracting path used for downsampling is defined by 5 convolution blocks, each with 2 (3 × 3) convolution layers (activation = “ReLU”, padding = “same”) and a (2 × 2) pooling layer. The UNET expansive path used for upsampling is defined by 5 convolution blocks, each with a (2 × 2) transpose convolution layer (activation = “ReLU”, padding = “same”), concatenation layer, and 2 (3 × 3) convolution layers (activation = “ReLU”, padding = “same”). A (1 × 1) output layer using a sigmoidal activation function returns the model output. Binary focal entropy is selected as the loss function due to large class imbalance between foreground and background pixels.

The model output is a pixelwise probability map for each input patch. The probability map values range from 0 to 1; values closer to 0 represent pixels more likely to belong to the negative class and pixels closer to 1 represent pixels more likely to belong to the positive class.

#### 2.4.4 Metrics for performance evaluation of U-Net model

We evaluate the performance of U-Net DL model in extracting each of the three lesions in terms of accuracy, recall, precision, F1-score, IoU and Dice score. The mathematical equations that describe each of these metrics are shown below:

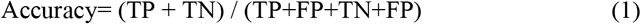

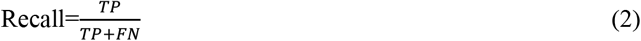

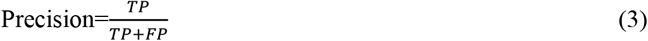

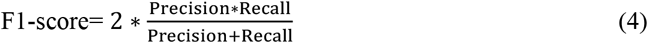

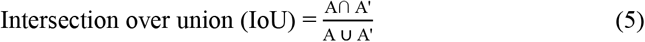

Where:

TP= True Positives (the sum of positive (foreground) pixels classified by the model)

TN= True Negatives (sum of correctly classified negative pixels)

FP= False Positives (the sum of negative (background) pixels misclassified by the model)

FN= False Negatives (the sum of misclassified positive pixels)

A = Area of ground truth pixels and A’
s = Area of predicted pixels

High recall values indicate that most of pixels belonging to the positive class (lesions) are predicted correctly by U-Net segmentation models. High precision values across the three lesion types also demonstrate the U-Net models successfully differentiate between lesion foreground and non-lesion background regions. High accuracy and F1 scores values reflect the excellent model performance and robustness. IoU is a useful metric for image segmentation by measuring the overlap between predicted and ground truth segmentation masks. This can be done for a class by dividing the intersection (overlap) of ground truth and predicted pixels belonging to the class by the total number of pixels in both masks belonging to the class. IoU score ranges from [0 – 1], where scores closer to 1 indicate greater agreement between predicted and ground truth masks.

### 2.5 Combination of TL and lesion features

The DR-detector framework performs a fusion of the TL features with lesion features to get a better representation of the features used for detection of DR. This is achieved by concatenating the flat representation of the features obtained by TL model (either VGG-16 or ResNet-50) with three lesion features that captures EXs, MAs and HEMs via image segmentation technique (Figure1C).

By combining the TL features and the lesion features, the resulting feature vector for each image in the training dataset that will be used for training the XGboost DR-detector model would be of size=25,088+3 =25,091 dim for VGG-16 and 100,352+3=100,355 dim for ResNet-50.

### 2.6 Metrics for performance evaluation of DR detection

The accuracy, precision, recall, F1-score and the area under the ROC curve (AUC) [58] have been used to evaluate the Xgboost model’s performance deployed in the DR detector framework. The mathematical equations that define each of the first four metrics were previously defined in equations [1–4] for evaluating the performance of U-Net segmentation models for lesion types. However, TP,FP,FN and TN terms for DR-detection task have a different indication from their previous annotation for evaluting the image segmentation task using U-Net model and hence are described below:

- TP refers to the number of correctly classified DR images.
- FP refers to the number of healthy images misclassified as DR images
- FN refers to the number of DR images misclassified as healthy images
- TN refers to the number of correctly classifed healthy images.

As for the AUC metric, it measures the entire two-dimensional area under the ROC curve [59] which measures how accurately the model can distinguish between DR and healthy images.

### 2.7 Experimental setup and model development

All experiments in this study were executed on Ubuntu Linux server with 128GB of RAM, 16 Intel Xeon E5-2609 1.7GHZ CPU cores, and 8 GPU cards. The optimized distributed gradient boosting python library [60] has been used for implementing the XGBoost model. The scikit-learn toolkit [61], the free machine learning python library has been used for implementing other ML models that were developed as a proof of concept to show that XGBoost was chosen because it outperforms other ML models (see Table 4 and 5). The Keras free python library [56] with tensorflow back end [57] was used to implement the TL models as well as the deep neural network (DNN) models to compare the performances in DR detection to the XGboost as an evidence to show the outperformance of later model.

## 3. Results

### 3.1 U-Net model performance evaluation

U-Net model performance evaluation results are shown in Table 3. The table shows an outperformance of the U-Net model for predicting MAs and EXs lesions over HEMs lesions. In other words, the general trend we observed was that the performance evaluation results for the HEMs segmentation model were consistently lower than those for the MAs and EXs models. We attribute this to the variation in the appearance of retinal hemorrhages, which can range from small, concentrated regions (dot hemorrhages) to larger and more irregular shapes. Examples of U-Net model predictions are shown in Supplementary Figure 3.

**Table 3:**
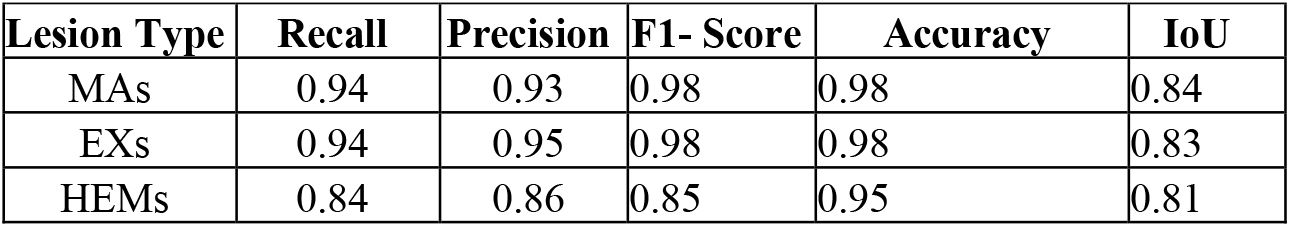
performance evaluation results of UNET model on extracting each type of lesions feature.

### 3.2 DR detection using VGG-16 and lesion features

As a proof of concept, we developed different ML models and trained them on the combination of VGG-16 and lesion features including the support vector machine (SVM), K-Nearest Neighbors (KNN), Xgboost, Logistic Regression (LR), Multi-Layered Perceptron (MLP), Decision Tree (DT), and Random Forest (RF) with default settings in addition to a DNN model with different structures including one input layer with 128 nodes, one input layer with 256 nodes and one hidden layer with 128 nodes, and one input layer with 512 nodes and 2 hidden layers with 256 and 128 nodes respectively. As shown in Table 4, XGBoost outperformed all other ML and DNN models achieving 99.38% accuracy in detecting DR.

**Table 4:** The performance of all Ml models and DNN for detection of DR using a combination of **VGG-16** and retinal lesion features

| Classifier | Accuracy % | Precision | Recall | F1-score | AUC |
| --- | --- | --- | --- | --- | --- |
| Xgboost | 99.38 | 1 | 0.99 | 0.99 | 0.994 |
| KNN | 47.53 | 0.45 | 0.25 | 0.32 | 0.475 |
| LR | 51.85 | 0.52 | 0.57 | 0.54 | 0.519 |
| SVM | 43.21 | 0.45 | 0.62 | 0.52 | 0.432 |
| MLP | 53.09 | 0.52 | 0.93 | 0.66 | 0.531 |
| DT | 88.27 | 1.00 | 0.77 | 0.87 | 0.883 |
| RF | 68.52 | 0.71 | 0.63 | 0.67 | 0.685 |
| **DNN (1 layer)** | 48 | 0.48 | 0.58 | 0.53 | 0.481 |
| **DNN (2 layers)** | 50 | 0.5 | 0.38 | 0.43 | 0.5 |
| **DNN (3 layers)** | 52 | 0.52 | 0.56 | 0.54 | 0.519 |

### 3.3 DR detection using ResNet-50 and lesion features

Table 5 shows the performance evaluation results of all DR detection models using the combination of ResNet-50 and lesion features. As can be observed from the table, XGBoost continued to outperform all other ML and DNN models achieving 100% accuracy for detection of DR.

**Table 5:** The performance of all Ml models and DNN for detection of DR using a combination of **ResNet-50** and retinal lesion features

| **Classifier** | **Accuracy %** | **Precision** | **Recall** | **F1-score** | **AUC** |
| --- | --- | --- | --- | --- | --- |
| **Xgboost** | **100** | **1.00** | **1.00** | **1.00** | **1.0** |
| **KNN** | 42.59 | 0.37 | 0.21 | 0.27 | 0.426 |
| **LR** | 53.09 | 0.52 | 0.68 | 0.59 | 0.531 |
| **SVM** | 45.68 | 0.48 | 0.84 | 0.61 | 0.457 |
| **MLP** | 47.53 | 0.49 | 0.95 | 0.64 | 0.475 |
| **DT** | 96.30 | 1 | 0.93 | 0.96 | 0.963 |
| **RF** | 56.79 | 0.55 | 0.70 | 0.62 | 0.568 |
| **1 layer-DNN** | 43.8 | 0.42 | 0.33 | 0.37 | 0.44 |
| **2 layer-DNN** | 48 | 0.49 | 0.75 | 0.59 | 0.481 |
| **3 layer-DNN** | 49 | 0.49 | 0.46 | 0.47 | 0.488 |

### 3.4 Performance of DR-detector with a single type of feature to understand feature importance and contribution

We decided to deploy XGBoost model in DR-detector framework for automatic detection of DR since it outperforms all other ML and DNN models as highlighted in the previous sections and documented in Tables 4 and 5. For deep analysis of the features that highly contribute to the performance of XGBoost model in the detection of DR, we have analyzed the importance of each feature by evaluating the XGBoost model performance with each type of extracted features including the TL and lesion features. This has been achieved by building three versions of the XGBoost model, where each version of the model is trained with one type of feature. Figure 2 shows a bar chart that represents the performance of XGBoost with VGG-16, ResNet-50 and lesions features (Supplementary Table 1). Clearly, we see a significant outperformance of the lesion features over the TL features that have been either extracted by VGG-16 or Resnt-50 models. This highlights the importance of the clinically manifested symptoms reflected in the form of lesion features in the detection of DR and how they can complement the textural features extracted by the TL model to improve the XGBoost performance and its interpretability for clinical use in detecting the presence of DR disease.

### 3.5 Performance of DR-detector with all possible feature combinations

Figure 3 shows a bar chart that represents the performance of XGBoost (DR-detector model) on the testing dataset with all possible combinations of TL and lesion features (Supplementary Table 2). Clearly, the figure shows that (Resent-50 and lesion) feature combination leads to the best performance of XGBoost model among all combinations and is slightly better than the performance of the model using (VGG-16 and lesions), and (VGG-16, ResNet-50, and lesions) combinations that lead to equal model performances. The figure also shows poor performance of XGBoost with VGG-16 and ResNet-50 combination (i.e., when the lesion features are specifically excluded) which also highlights the importance of lesion features in the detection of DR.

**Figure 3.**
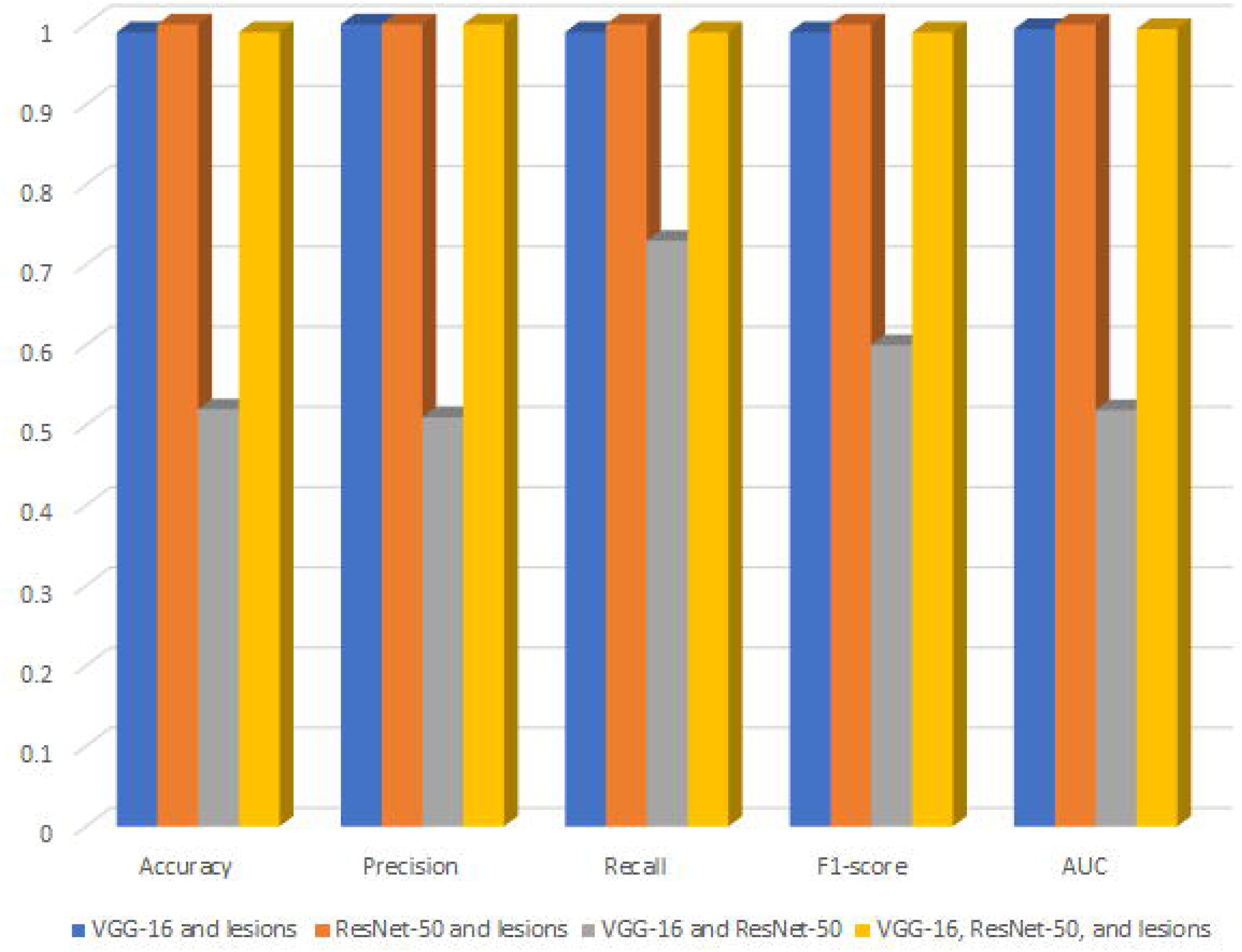
The performance of XGBoost with all possible combinations of TL and lesion features.

### 3.6 Lesion feature importance and their effect on XGBoost performance

For deep analysis of the contribution of each lesion feature to the performance of DR-detector model in DR detection, we have reported the importance of each lesion feature through training and testing the XGBoost model with each type of lesion feature individually. This has been achieved by building three versions of the XGBoost model, where each version of the model is trained with one lesion feature. Figure 4 shows the performance of XGBoost with EXs, MAs and HEMs lesion features as well as with their combinations (Supplementary Table 3). Clearly, the table shows a significant outperformance of XGBoost model using HEMs lesion feature over its performance using either the MAs or EXs lesion features which equally contribute to the performance of the model. More importantly, the model also achieves better performance using HEMs alone than its performance using the combination of the three lesion features altogether. Thus, our results highlight the importance of HEMs as a feature in the detection of DR and how it can be used to allow the interpretability of the model for clinical use in DR detection.

**Figure 4.**
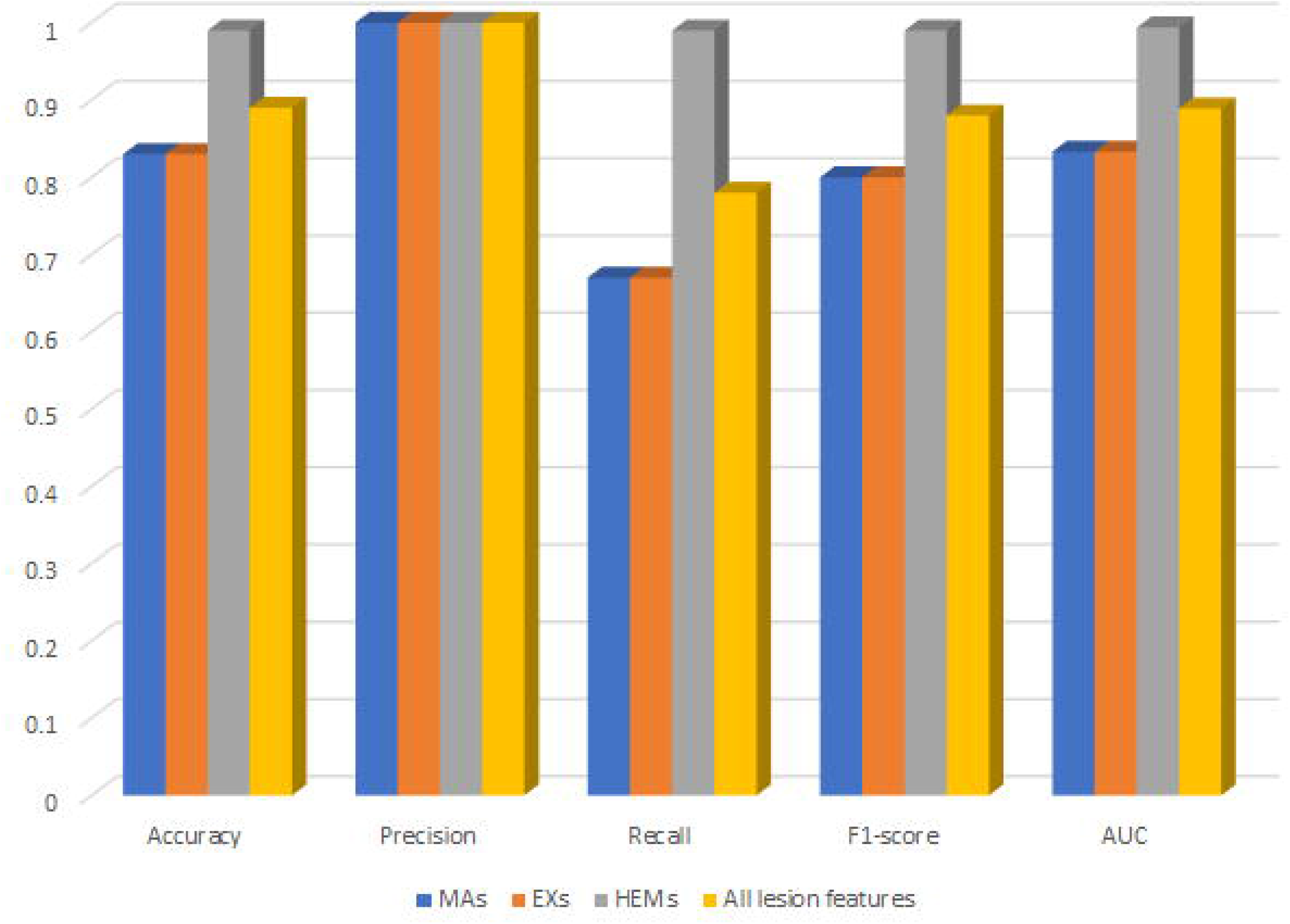
The performance of XGBoost with each type of lesion features as well as their combinations.

## 4. Discussion

There are several observations that can be summarized from our experimental results. First, using our proposed approach we found that the integration of lesion features with the TL features significantly improves the performance of the DR-detector model and adds a clear importance to its clinical interpretability. However, currently it is not possible to only work with the lesion features for training the DR-detector model as there are few retinal imaging datasets in the literature that provide the image masks corresponding to the retinal images in the dataset that are needed to extract and quantify the lesion features using the U-Net segmentation models and obtaining those masks need the involvement of trained ophthalmologists which is costly and time-consuming.

It was also observed that deploying the XGBoost model as an ensemble of ML classifiers in the DR-detector framework led to a better binary classification of DR and error detection than deploying dense classifiers of DL models as introduced in few previous studies [20, 37,38]. This motivated us to use the flat combination of TL and lesion features (Figure 1C) which is simpler, straight forward and more convenient to be applied to the XGBoost ML model than combining both types of features using different pooling approaches [20], or by extending the last fully connected (FC) layer of TL model with additional number of neurons equal to the number of lesion features and using softmax function to obtain the predictive DR class probabilities [37].

Regarding the contribution of lesion features to the DR-detector (XGBoost) model performance, our approach shows that lesion features have different contributions to the model performance. Particularly, it shows a significant contribution of the hemorrhages over the other two lesion features for binary identification of DR (Supplementary Table 3). This is likely because hemorrhages may arise early during the progression of DR and are associated with the worsening of vision and the development of other vision-threatening lesions. Our observations highlight the importance of such a feature to train the binary classifier of DR-detector framework and adds value for its clinical usage in DR diagnosis. However, it remains to be seen how our presented results will hold when testing our lesion feature-based models on different fundus retinal testing datasets with more complex demographics and varying quality.

It is also noteworthy to mention that although we have applied the DR-detector framework to a relatively small training and testing datasets in comparison with the large datasets that have been used in the literature for DR detection (e.g.,[17],) our datasets are from heterogenous sources i.e., have different variety of retinal images that were imported from multiple publicly available datasets with different settings of capturing the fundus retinal images. This, of course, highlights the efficiency of our proposed framework in detecting the initial signs of DR, even when applied to a set of retinal fundus images that were not imported from the same resource. Thus, we expect a better performance of DR-detector framework with larger training and testing datasets in the future.

## 5. Conclusions and Future work

In this study, we have proposed a framework called DR-detector that combines the features extracted from fundus retinal image by transfer learning model and the lesion features extracted using semantic image segmentation via U-Net DL model for accurate detection of DR using the XGboost ML model deployed in this framework. The model was trained using the combination of both features on a training dataset collected from various publicly available datasets and was tested on an external dataset that consists of DR images from IDRID dataset and healthy images from HRF and Messior-2 dataset. Our experimental results show that our proposed framework for DR detection achieves a testing accuracy of 100% in detecting DR using the combination of Resnt-50 and lesion features and 99% testing accuracy using the combination of VGG-16 and lesion features. Based on these results, we arrived at the conclusion that the extracted clinically relevant lesion features have a significant impact on the performance of the DR-detector model and would be an excellent complement to the textural features extracted by the TL model to improve the performance of DR-detector model and its interpretability for clinical use for detecting the presence of DR disease.

We anticipate a natural extension of our current work is to first extend to other TL models that are commonly used in the literature to study how they perform for DR classification task and then expand our general framework to be applied for the prediction of different severity levels of DR. Finally, we are looking forward to applying our approach to combine TL features with other types of DR lesion features such as cotton wool spots, foveal avascular zone, optic disc, and retinal blood vessels since these features were known in the literature to be associated with greater severity of DR [62]. This might lead to better performance of the DR-detector model and its interpretability for clinical use on much larger and clinically diverse datasets, especially if it will be extended to predict the various grades of DR.

## Data Availability

All data produced are available online at:
1. https://www.adcis.net/en/third-party/e-ophtha/
2. https://github.com/WeiQijie/retinal-lesions
3. https://www.kaggle.com/c/aptos2019-blindness-detection/data
4. https://www5.cs.fau.de/research/data/fundus-images/
5. https://idrid.grand-challenge.org
6. https://www.adcis.net/en/third-party/messidor2/

https://github.com/Janga-Lab/DR-detector

https://www.adcis.net/en/third-party/e-ophtha/

https://github.com/WeiQijie/retinal-lesions

https://www.kaggle.com/c/aptos2019-blindness-detection/data

https://www5.cs.fau.de/research/data/fundus-images/

https://idrid.grand-challenge.org

https://www.adcis.net/en/third-party/messidor2/

## Author Contributions

DH, HMG, MH, ADB, AH and SCJ conceived and designed the study. DH did the TL feature extraction using VGG-16 and ResNet-50 models. HMG did the lesion feature extraction using image segmentation via U-Net model. DH combines TL and lesion features for training and evaluating the XGboost model of DR-detector framework. DH and HMG wrote the manuscript. DH and HMG contribute to the DR-detector GitHub repository. ADB, AH and SCJ revised the manuscript and provided valuable comments.

## Acknowledgement

This work is supported by the IUPUI Institute of Integrative Artificial Intelligence (iAI).

## Footnotes

1 https://www.kaggle.com/c/aptos2019-blindness-detection/data

