## Supplementary Tables for "Combining Transfer Learning with Retinal Lesions Features for Accurate Detection of Diabetic Retinopathy"

| **Type of features** | Accuracy | Precision | Recall | F1-score | AUC |
| --- | --- | --- | --- | --- | --- |
| **VGG-16 features** | 46.30 | 0.47 | 0.57 | 0.51 | 0.463 |
| **ResNet-50 features** | 49.38 | 0.49 | 0.49 | 0.49 | 0.494 |
| **Lesions features** | **88.89** | **1.0** | **0.78** | **0.88** | **0.889** |

**Supplementary Table 1.** The performance of XGboost on the testing dataset with each type of features (Vgg-16, ResNet-50, and lesion features).

| **Features combination** | Accuracy | Precision | Recall | F1-score | AUC |
| --- | --- | --- | --- | --- | --- |
| **VGG-16 and lesions** | 99.38 | 1 | 0.99 | 0.99 | 0.994 |
| **ResNet-50 and lesions** | **100** | **1.00** | **1.00** | **1.00** | **1.0** |
| **VGG-16 and ResNet-50** | 51.85 | 0.51 | 0.73 | 0.6 | 0.519 |
| **VGG-16, ResNet-50, and lesions** | 99.38 | 1 | 0.99 | 0.99 | 0.994 |

**Supplementary Table 2** The performance of XGboost on the testing dataset with all possible combinations of TL and lesion features.

| **Type of features** | Accuracy | Precision | Recall | F1-score | AUC |
| --- | --- | --- | --- | --- | --- |
| MA | 83 | 1.00 | 0.67 | 0.8 | 0.833 |
| EX | 83 | 1.00 | 0.67 | 0.8 | 0.833 |
| HEM | **99.38** | **1:00** | **0.99** | **0.99** | **0.994** |
| All lesion features | 88.89 | 1:00 | 0.78 | 0.88 | 0.889 |

**Supplementary Table 3.** The performance of the DR-detector model (XGBoost) on the testing dataset with each type of lesion features (MA,EX, and HEM).
