## Supplementary Figures for "Combining Transfer Learning with Retinal Lesions Features for Accurate Detection of Diabetic Retinopathy"

Supp_Figure 1

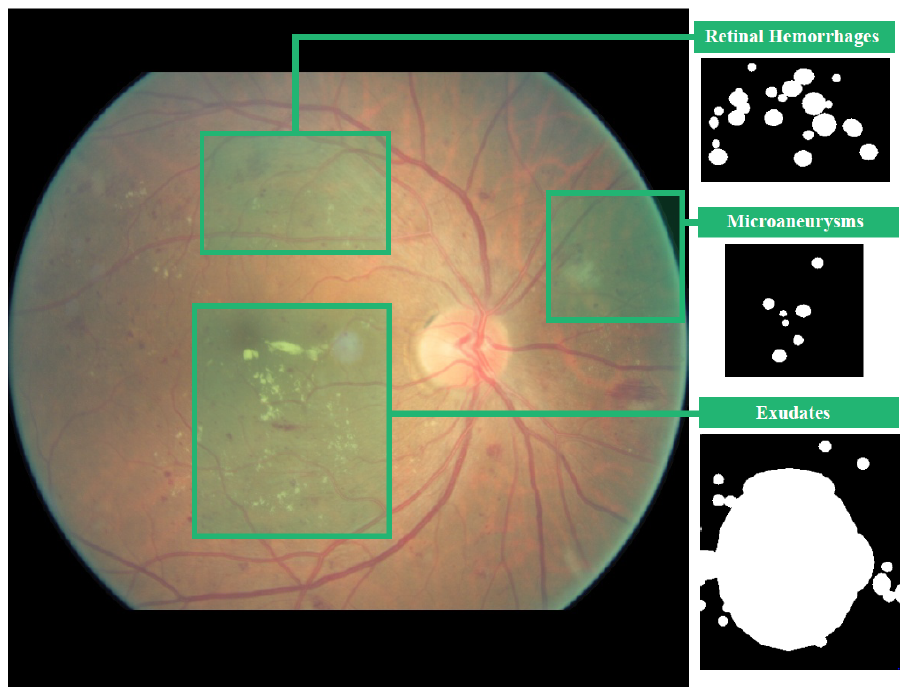

Supp_Figure 2

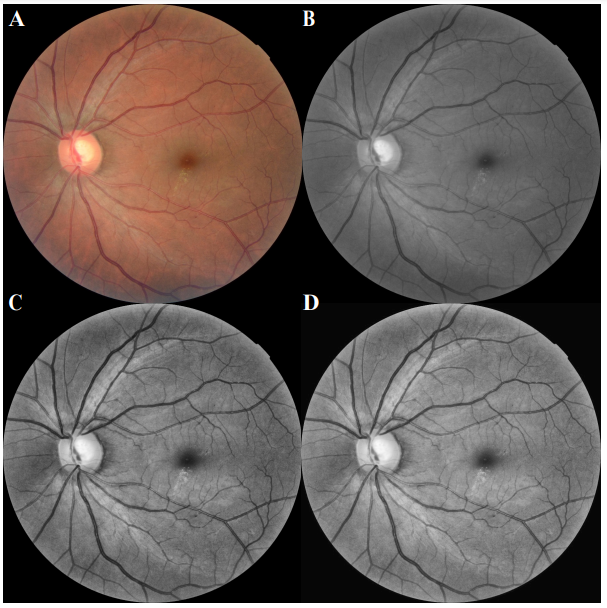

**Supp_Figure 3**

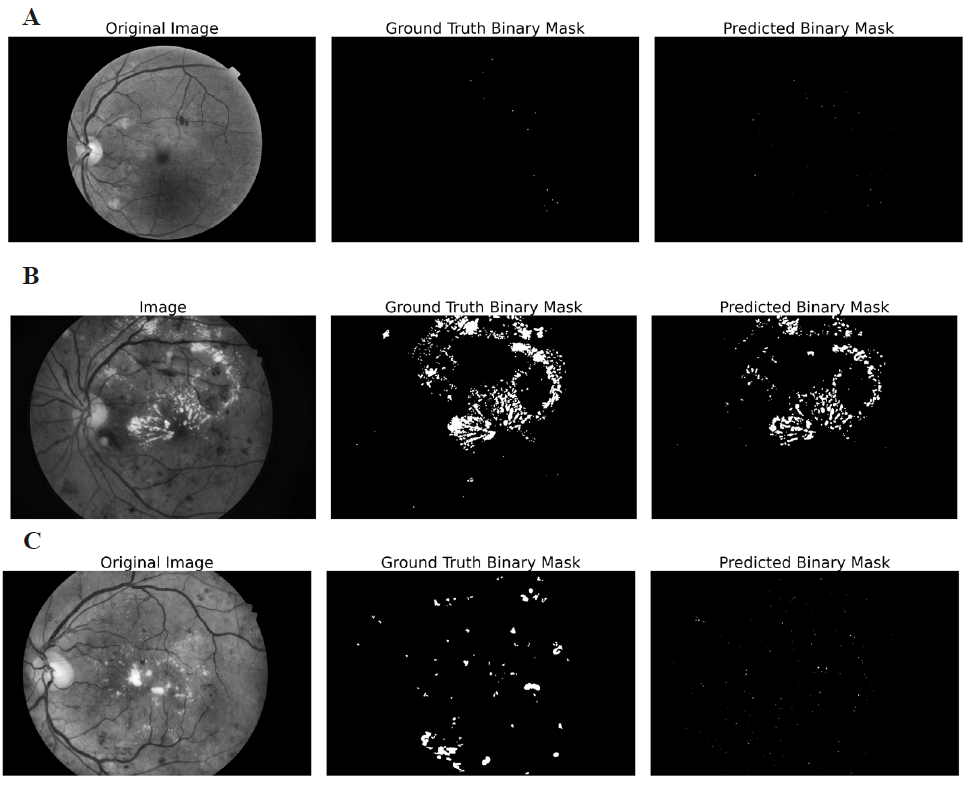

A= An example UNET model predictions of Microaneurysms

B= UNET model predictions of Exudates

C= UNET model predictions of Hemorrhages
